# Genetic and Environmental Predictors of Seasonality and Seasonal Affective Disorder in Individuals with Depression

**DOI:** 10.64898/2026.04.22.26351539

**Authors:** Floris Huider, Jacob J. Crouse, Sarah E. Medland, Ian B. Hickie, Nicholas G. Martin, Jodi T. Thomas, Brittany L. Mitchell

## Abstract

**Background:** The etiology and nosological status of seasonal affective disorder (SAD) as a specifier of depressive episodes versus a transdiagnostic disorder are the subject of debate. In this study, we investigated the underlying etiology of SAD and dimensional seasonality by examining their association with latitude and genetic risk for a range of traits, and investigated gene-environment interactions.

**Methods:** This study included 12,460 adults aged 18-90 with a history of depression from the Australian Genetics of Depression Study. Regression models included predictors for latitude (distance from equator) and polygenic scores for eight traits; major depressive disorder, bipolar disorder, anxiety disorders, chronotype, sleep duration, body mass index, vitamin D levels, and educational attainment. Outcomes were SAD status and general seasonality score.

**Results:** SAD was positively associated with latitude (OR[95%CI] = 1.05[1.03-1.06], p_adjusted_<0.001), and there was nominal evidence of additive and multiplicative interactions between chronotype genetic risk and latitude (OR = 0.99[0.99-0.99], p_adjusted_=0.381; OR=0.98[0.97-0.99], p_adjusted_=0.489). General seasonality score was associated with latitude (IRR=1.01[1.01-1.01], p_adjusted_ 0.001) and genetic risk for major depressive disorder (IRR =1.02[1.01-1.03], p_adjusted_<0.001), bipolar disorder (IRR=1.02[1.01-1.03], p_adjusted_=0.001), anxiety disorders (IRR=1.03[1.01-1.04], p_adjusted_<0.001), vitamin D levels (OR=0.89[0.80-0.95], p_adjusted_=0.048), and educational attainment (IRR=0.97[0.96-0.99], p_adjusted_<0.001).

**Conclusions:** These findings enhance understanding of SAD etiology, highlighting contributions of psychiatric genetic risk and geographic measures on seasonal behavior, and support examining seasonality as a continuous dimension.

## Introduction

For most people, behavior and mood tend to change with the passing of seasons (1,2). Seasonal affective disorder (SAD) can be characterized as the clinical extreme of this phenomenon (3,4). Prevalence of SAD varies by both definition and region (1 – 16%), with higher prevalences observed for broader criteria, such as subsyndromal SAD, and in regions at higher latitudes (further away from the equator) (5,6). SAD is recognized by many as a major mental health concern and public health issue. It has a highly recurring pattern (7) and causes substantial impairments (8), including notable socio-economic consequences, greater severity (in bipolar disorder), and heightened cardiovascular risk. Recognizing its widespread impact, an expert consensus by the International Society for Bipolar Disorders Chronobiology and Chronotherapy Task Force emphasizes seasonality as a critical factor in clinical practice (9).

The concept of SAD was introduced four decades ago and has been revised over the years. SAD encompasses depressive symptoms (low mood, changes in sleep, activity, and appetite patterns) with typical onset during autumn-winter and remission in spring-summer. The fifth edition of the Diagnostic and Statistical Manual of Mental Disorders (DSM-5) defines SAD as a specifier of unipolar or bipolar depressive episodes. As a specifier, SAD contributes to 22-25% of depression cases (10,11). SAD is also commonly recognized as a transdiagnostic disorder or dimensional continuum, in that seasonality symptoms cross clinical boundaries and are normally distributed in the general population (12,13). The general seasonality score (GSS) is a common seasonality measure that captures the dimensional aspect of seasonal symptom variation, offering a transdiagnostic measure that reflects the broader continuum of seasonality in the population. Examining seasonality both as a dichotomous diagnosis (SAD) and as a continuous variable (GSS) can provide valuable insights into the clinical nosology of seasonality.

Several risk factors for SAD have been proposed, including genetic predisposition, chronotype, latitude and changes in light exposure (14). With a heritability of up to 29% (15) across its behavioral dimensions, there is a substantial genetic component to SAD, one that is similar in size to that of major depressive disorder (35%) (16) but lower than bipolar disorder (70-90%) (17). Environmental risk factors mainly center around photoperiod (day length) and light exposure, which have been shown to affect physiological circadian and infradian (e.g., seasonal) cycles, gene expression, and sleep (14). An association between light exposure and affective disorders is further corroborated by light therapy being the recommended first-line treatment approach for SAD (18). Latitude is a common proxy for photoperiod and has been positively associated with SAD prevalence (5). However, this association does not always translate across populations (19,20). Importantly, SAD is only rarely studied in the Southern hemisphere (19), despite photoperiod being highly dependent on hemisphere and further fine-tuned geographical location.

The true etiology of SAD is likely a complex and multifactorial interaction of genetic and environmental risk factors (6,21), as is the dominant theory for many mental health disorders (diathesis-stress model) (22). Gene-environment interaction (GxE) can be a substantial contributor to individual differences in complex traits (23,24), as is the case for, e.g., major depressive disorder and trauma (25). GxE has also been proposed as a potential contributor to the heterogeneity in affective disorders (26) and inconsistency of risk factors in seasonal pattern findings (27). For example, an increased genetic vulnerability to depression or seasonal changes may be more likely to manifest at higher latitudes. Yet, to our knowledge, the interaction between genetic risk and latitude on SAD has not been studied.

Recent advances in the field of molecular genetics and increased sample sizes (and associated statistical power of gene-finding studies) allow for genome-wide interrogation of associations with complex traits (28), and the derivation of genetic risk indices, termed polygenic scores. These polygenic scores reflect an individual’s genetic predisposition for a trait based on their entire allelic makeup and allow for a better approximation of genetic risk than any singular gene or variant is likely to provide (29).

In this paper, we examine whether latitude and genetic risk for three psychiatric traits—major depressive disorder, bipolar disorder, and anxiety disorders—and five correlates of seasonality— chronotype, sleep duration, body mass index, vitamin D levels, and educational attainment—explain variation in SAD. Specifically, we examine whether latitude and polygenic scores for these traits are associated with SAD and whether genetic risk and latitude interact to predict SAD. Furthermore, we extend these analyses to examine seasonality as a continuous dimension, contributing to the ongoing debate on its classification within psychiatric nosology. We approach this question using the Australian Genetics of Depression Study (N = 12,460), enabling examination of differential vulnerability within an at-risk group.

## Methods and Materials

### Sample characteristics

This study used data from the Australian Genetics of Depression Study (AGDS). The AGDS is a nation-wide initiative aimed at studying the genetic and environmental risk factors for depression (30) and it comprises over 22,000 Australian adults sampled from the general population, who responded to a media appeal seeking participants who had been diagnosed with depression or had been prescribed antidepressants in the five years prior to enrollment. Individuals were included in the current study if they were of European ancestry and had non-missing data on seasonality, age at measurement, and genotype information (N = 12,460). All relevant ethical regulations for working with human participants were followed in the conduct of the study, and written informed consent was obtained from all participants. All protocols and questionnaires were approved by QIMR Berghofer Medical Research Institute Human Research Ethics Committee in Brisbane, Australia (P2118).

### Measures

#### Seasonality

Seasonality data were derived from the 17-item self-report Seasonal Pattern Assessment Questionnaire (SPAQ; see Supplementary Text, Psychometric information for the SPAQ) (12,31). We considered six items measuring the degree of typical seasonal change in sleep length, social activity, mood, weight, appetite, and energy, on a scale from 0 (no change) to 4 (extremely marked change). Item scores were summed to derive the general seasonality score (GSS; 0 – 24). Participants were asked to indicate the month(s) in which they tend to feel worst (if any) and to report whether symptoms were a problem for them, and to which degree (mild, moderate, marked, severe, disabling).

Seasonal affective disorder (SAD) cases were identified as having a GSS ≥ 11, symptoms of at least a moderate severity, and feeling worst during Australian summer (December, January, February) and/or winter (June, July, August) (Figure S1). This definition reflects typical seasonal delineations in Australia and slightly deviates from the original definition by Kasper et al. (1), which was developed in the North American context and considers only January or February and July or August for the ‘feel worst’ calendar item. SAD ‘controls’ were defined as participants who answered all questions but did not meet the criteria for being a SAD ‘case’ (these remaining participants having other forms of depressive disorders).

#### Polygenic scores

Polygenic scores (PGS) reflect an individual’s genetic predisposition for a trait of interest, and are calculated by summing the number of risk-increasing and protective genetic variants, weighted by results from a well-powered genome-wide association study (GWAS) (29). We calculated PGS for eight traits using publicly available data as listed in Table 1. PGS weights were calculated in SBayesR (32) (see Supplementary Text, Genotype information and polygenic score generation). All PGS were standardized so that effect sizes reflected the change in the outcome per standard deviation increase in PGS.

**Table 1.**
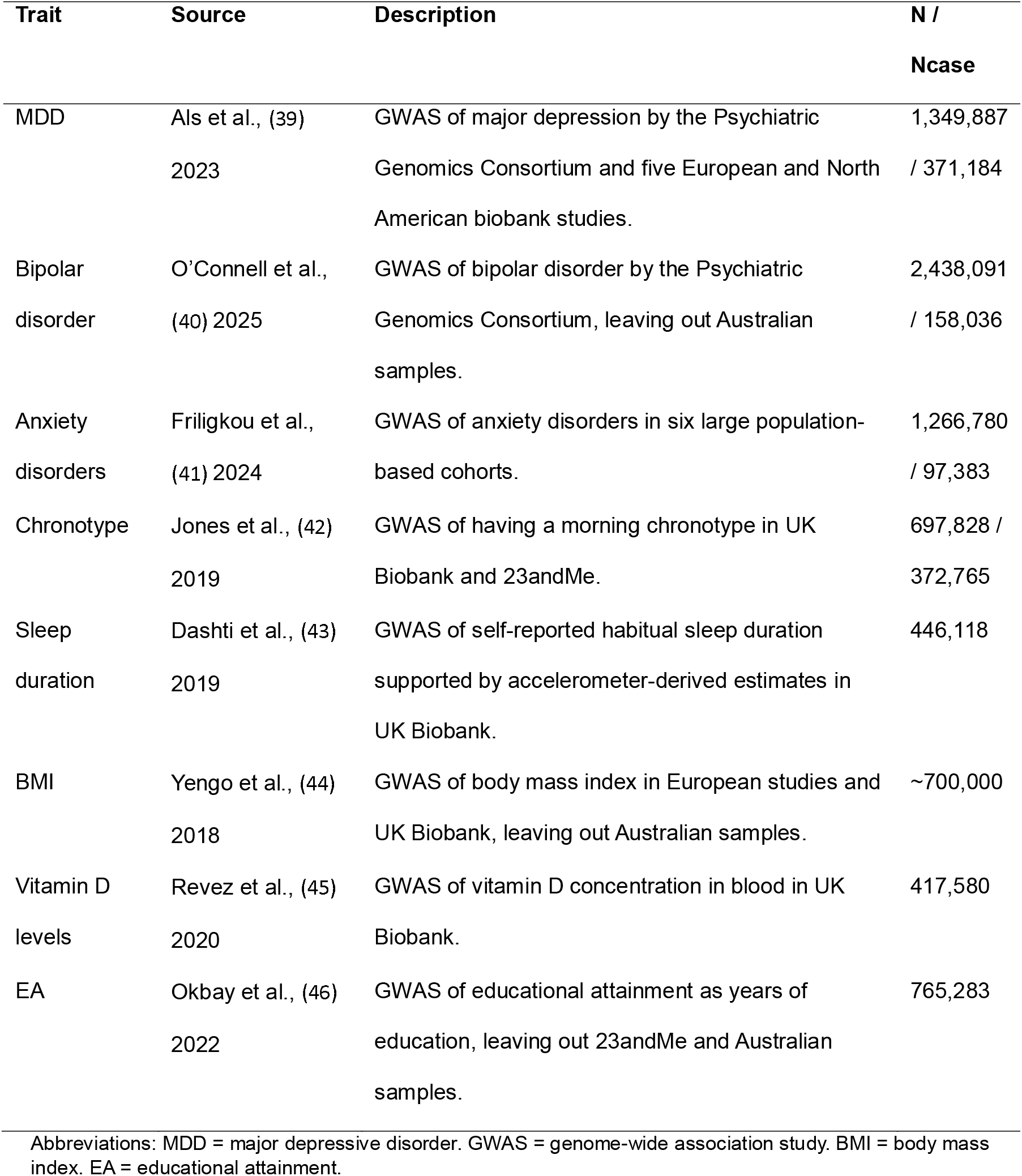
Discovery GWAS’ used to generate polygenic scores.

| Trait | Source | Description | N /<br>Ncase |
| --- | --- | --- | --- |
| MDD | Als et al., (39)<br>2023 | GWAS of major depression by the Psychiatric Genomics Consortium and five European and North American biobank studies. | 1,349,887<br>/ 371,184 |
| Bipolar disorder | O'Connell et al.,<br>(40) 2025 | GWAS of bipolar disorder by the Psychiatric Genomics Consortium, leaving out Australian samples. | 2,438,091<br>/ 158,036 |
| Anxiety disorders | Friligkou et al.,<br>(41) 2024 | GWAS of anxiety disorders in six large population-based cohorts. | 1,266,780<br>/ 97,383 |
| Chronotype | Jones et al., (42)<br>2019 | GWAS of having a morning chronotype in UK Biobank and 23andMe. | 697,828 /<br>372,765 |
| Sleep duration | Dashti et al., (43)<br>2019 | GWAS of self-reported habitual sleep duration supported by accelerometer-derived estimates in UK Biobank. | 446,118 |
| BMI | Yengo et al., (44)<br>2018 | GWAS of body mass index in European studies and UK Biobank, leaving out Australian samples. | ~700,000 |
| Vitamin D levels | Revez et al., (45)<br>2020 | GWAS of vitamin D concentration in blood in UK Biobank. | 417,580 |
| EA | Okbay et al., (46)<br>2022 | GWAS of educational attainment as years of education, leaving out 23andMe and Australian samples. | 765,283 |

Abbreviations: MDD = major depressive disorder. GWAS = genome-wide association study. BMI = body mass index. EA = educational attainment.

#### Latitude

Latitude was defined as distance from the equator based on the median latitude (as estimated from Google Maps API (https://developers.google.com/maps)) of the participant’s post code (zip code) recorded from demographic information collected at the time of recruitment. Latitude estimates, being initially negative for the southern hemisphere, were converted to positive values to aid interpretation, so that higher latitudes reflect a larger distance from the equator.

### Statistical Analyses

All analyses were conducted in R v4.4.0 (33), and all models included sex (inferred from genotype data), age, and 10 genetic ancestry principal components as covariates. A series of logistic regressions were used to test the associations between PGS and latitude with SAD as the outcome, with estimates reported as odds ratios (ORs). Two-component mixture zero-inflated negative binomial regression models were used to test the associations between PGS and latitude with GSS as the outcome (see Supplementary text, Model selection for general seasonality score and Figure S2 for the GSS distribution). The two-component mixture zero-inflated negative binomial regression model has two components: 1) a count model testing the association between the predictor and the non-zero GSS values, and 2) a zero-inflated model that tests the association between a predictor and the probability of having no seasonality symptoms (GSS = 0). For these two-component mixture zero-inflated negative binomial models with GSS as the outcome, estimates from the count model component are presented in the main text as incident rate ratios (IRRs), representing relative differences in expected symptom counts among individuals in the count-generating process. Estimates from the zero-inflation component are reported as odds ratios (ORs) in the Supplemental Information.

We first assessed the association of each polygenic score with SAD and GSS in separate single-PGS models. We then evaluated all eight polygenic scores together in the same multiple-PGS model. The main effect of latitude on SAD and GSS was assessed in a model with latitude as the single predictor. Next, latitude was included in the single- and multiple-PGS models. We then evaluated polygenic score × latitude interaction effects on both the additive and multiplicative scales. Additive interaction was tested using linear regression models, in which the product term represented deviation from additivity on the raw outcome scale. Multiplicative interaction was tested using logistic regression for SAD and zero-inflated negative binomial regression for GSS, in which the product term represented deviation from multiplicativity on the log-odds and log-count scales, respectively. All interaction models additionally included covariate main effects and covariate × polygenic score and covariate × latitude interaction terms to reduce confounding of the interaction estimate (34). P-values were corrected for multiple comparisons using the Benjamini-Hochberg procedure, controlling the false discovery rate at 5% across 34 tests, for SAD and GSS outcome variables separately.

#### Sensitivity analyses

In a series of sensitivity analyses we examined the robustness of models to different variable and model specifications. We assessed whether the association between seasonality and MDD-PGS depended on MDD severity, as defined by number of depressive episodes, symptoms, or duration. We also tested whether associations changed when using alternative definitions of SAD; following the example of Kasper et al. (1) where only two months of winter and summer were considered in determining SAD case-control status (rather than three as used in our main analyses), and where the ‘feel worst’ calendar criterion was dropped (see Supplementary Text, Sensitivity analyses methods).

## Results

### Sample characteristics

A total of 12,460 Australian Genetics of Depression Study (AGDS) participants were included for analyses of seasonal affective disorder (SAD) case-control status and the general seasonality score (GSS) (76% female; mean age = 44.1 years; case = 8.7%; Table 2, Figure S3). There were clear peaks in prevalence of feeling worst during the winter and summer months (Figure S4), providing support for including all three months of the meteorological seasons in determining SAD case-control status.

**Table 2.**
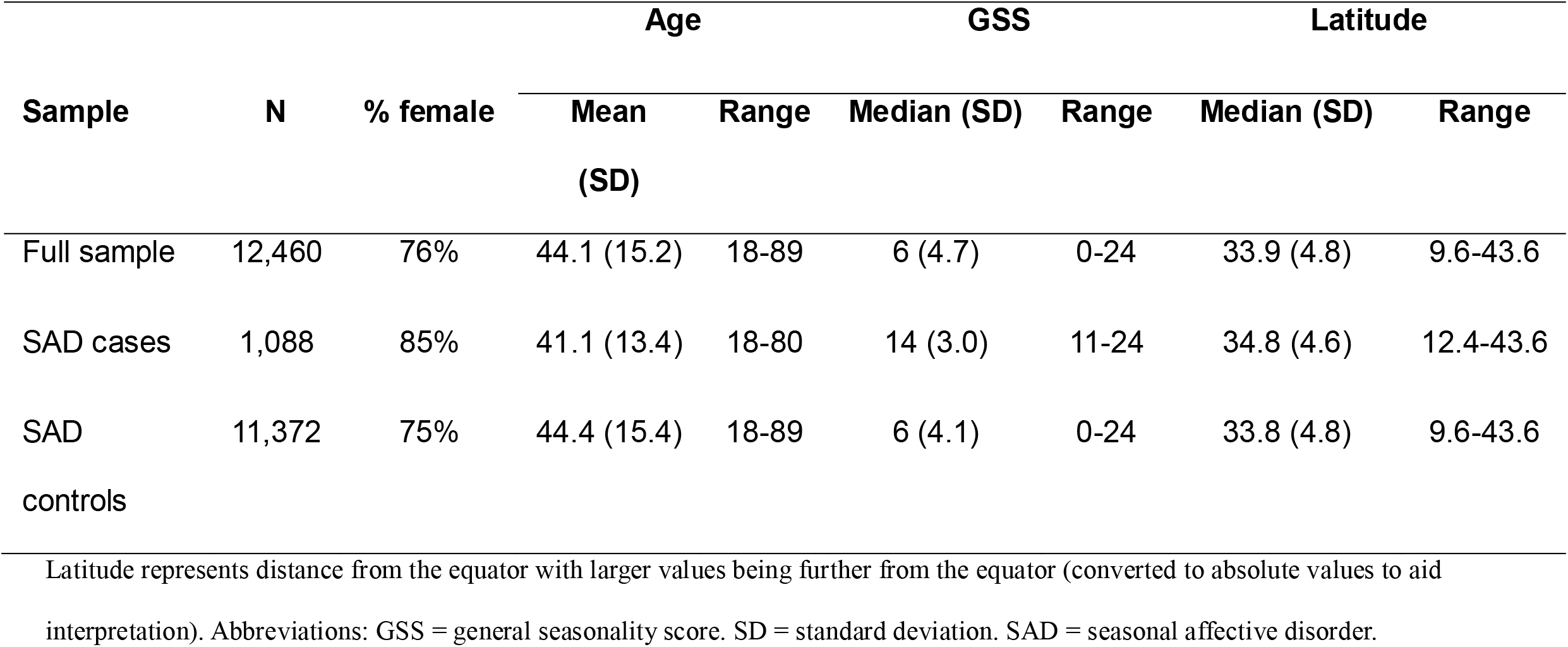
Sample characteristics.

| Sample | N | % female | Age |  | GSS |  | Latitude |  |
| --- | --- | --- | --- | --- | --- | --- | --- | --- |
|  |  |  | Mean<br>(SD) | Range | Median (SD) | Range | Median (SD) | Range |
| Full sample | 12,460 | 76% | 44.1 (15.2) | 18-89 | 6 (4.7) | 0-24 | 33.9 (4.8) | 9.6-43.6 |
| SAD cases | 1,088 | 85% | 41.1 (13.4) | 18-80 | 14 (3.0) | 11-24 | 34.8 (4.6) | 12.4-43.6 |
| SAD controls | 11,372 | 75% | 44.4 (15.4) | 18-89 | 6 (4.1) | 0-24 | 33.8 (4.8) | 9.6-43.6 |

Latitude represents distance from the equator with larger values being further from the equator (converted to absolute values to aid interpretation). Abbreviations: GSS = general seasonality score. SD = standard deviation. SAD = seasonal affective disorder.

### Genetic Risk and Seasonality

None of the polygenic scores (PGS) were significantly associated with SAD (Figure 1, Table S1). Genetic risk for major depressive disorder (MDD) and anxiety disorders were positively associated with SAD but did not survive multiple testing correction (MDD: OR[95% confidence interval (CI)]=1.08[1.01-1.15], p_adjusted_=0.124; anxiety disorders: OR=1.07[1.00-1.14], p_adjusted_=0.193). We found significant positive associations between the non-zero GSS values and genetic risk for MDD (Incidence Rate Ratio (IRR)[95%CI]=1.03[1.02-1.04], p_adjusted_<0.001), bipolar disorder (BD) (IRR=1.02[1.01-1.03], p_adjusted_=0.001), and anxiety disorders (IRR=1.03[1.01-1.04], p_adjusted_<0.001). Conversely, among individuals with any seasonality symptoms, genetic predisposition for higher educational attainment was significantly associated with a lower GSS (IRR=0.97[0.96-0.98], p_adjusted_<0.001). Genetic risk for vitamin D was not associated with non-zero GSS values, but was significantly negatively associated with GSS in the zero-inflated part of the model (OR = 0.89[0.82-0.96], p_adjusted_=0.048); an increasing genetic propensity for higher vitamin D concentrations was associated with a lower probability of having no seasonality symptoms at all (Table S1). No other PGS showed a significant association with GSS.

**Figure 1.**
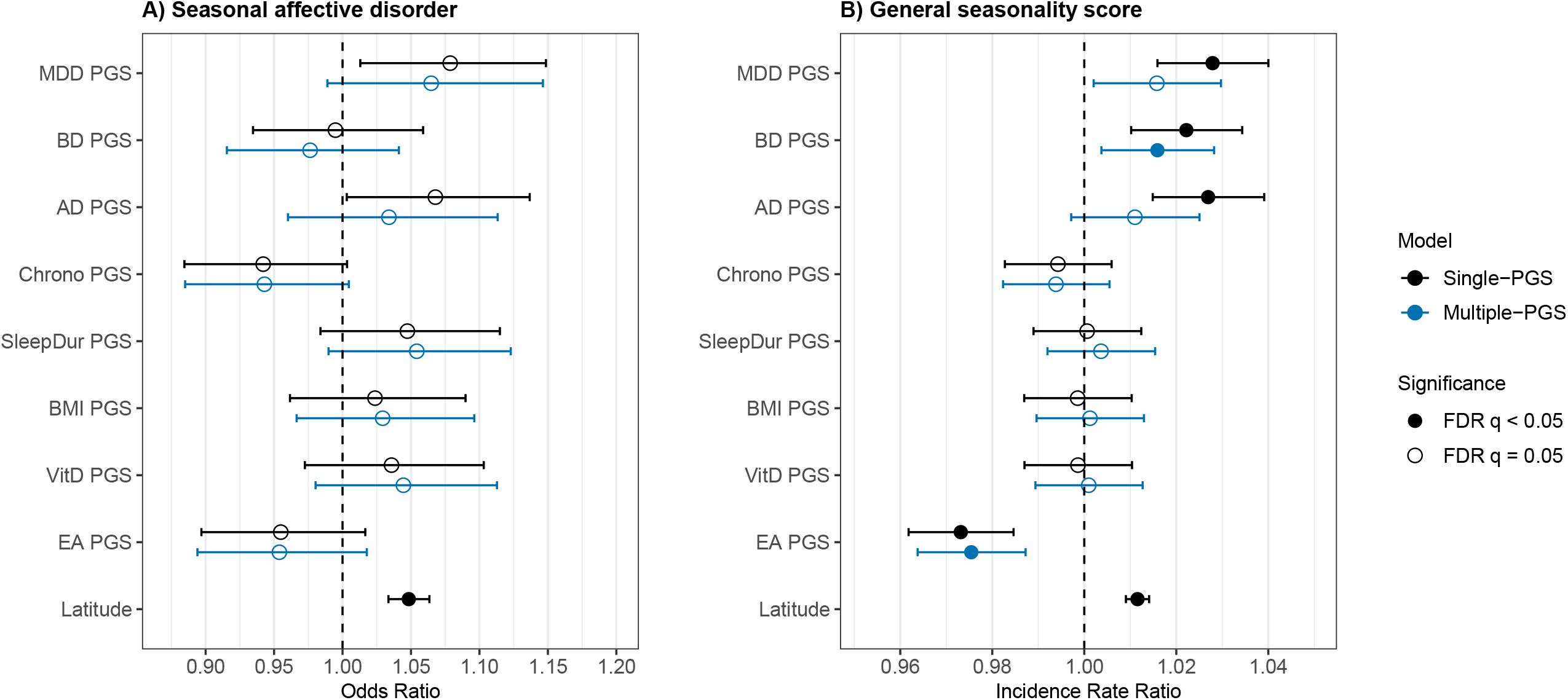
Association of polygenic scores and latitude with seasonality. Associations between polygenic scores and latitude on seasonal affective disorder (SAD) and general seasonality score (GSS) from single-PGS models and a multiple-PGS regression model including all polygenic score predictors. A) Main effects on SAD (N = 12,460). B) Main effects on non-zero GSS values (count component of the model) (N = 12,460). Note that the main effect results from the zero-inflated model that tests the association with the probability of having no seasonality symptoms (GSS = 0) can be seen in Table S1. Error bars represent 95% confidence intervals. A filled in circle indicates significance after multiple testing correction (Benjamini-Hochberg False Discovery Rate P < 0.05). Abbreviations: MDD = major depressive disorder. PGS = polygenic score. BD = bipolar disorder. AD = anxiety disorders. Chrono = chronotype. SleepDur = sleep duration. BMI = body mass index. VitD = vitamin D. EA = educational attainment.

Next, we included the PGS for all traits in one model predicting SAD and GSS. As a general trend, effects in this multiple-PGS model were very similar to the single-PGS models. However, the association of the MDD-PGS and anxiety disorders-PGS with non-zero GSS values were attenuated and became non-significant in the multiple-PGS model (Figure 1, Table S1). Lack of significance in the multiple-PGS model suggests that the effect of an individual PGS is not independent of other PGS, pointing to shared genetic pathways.

### Latitude and Seasonality

With increasing latitude, i.e., distance from the equator, there was a significant increase in SAD risk (OR=1.05[1.03-1.06], p_adjusted_<0.001) and non-zero GSS values (IRR=1.01[1.01-1.01], p_adjusted_<0.001) (Figure 1, Table S1). Adding latitude to the single-PGS models left genetic risk associations with SAD and GSS largely unattenuated (Table S2). There were no significant PGS-by-latitude interaction effects on SAD or GSS (Table S2). We found additive (β=-0.001[-0.002-0.0003], p=0.011, p_adjusted_=0.122) and multiplicative (OR=0.98[0.97-1.0, p=0.014, p_adjusted_=0.122) chronotype PGS-by-latitude interactions with SAD, however they did not survive multiple testing correction (Figure 2). The direction of effect suggests that latitude may have a smaller effect on SAD risk in individuals with a higher genetic propensity for morning chronotype.

**Figure 2.**
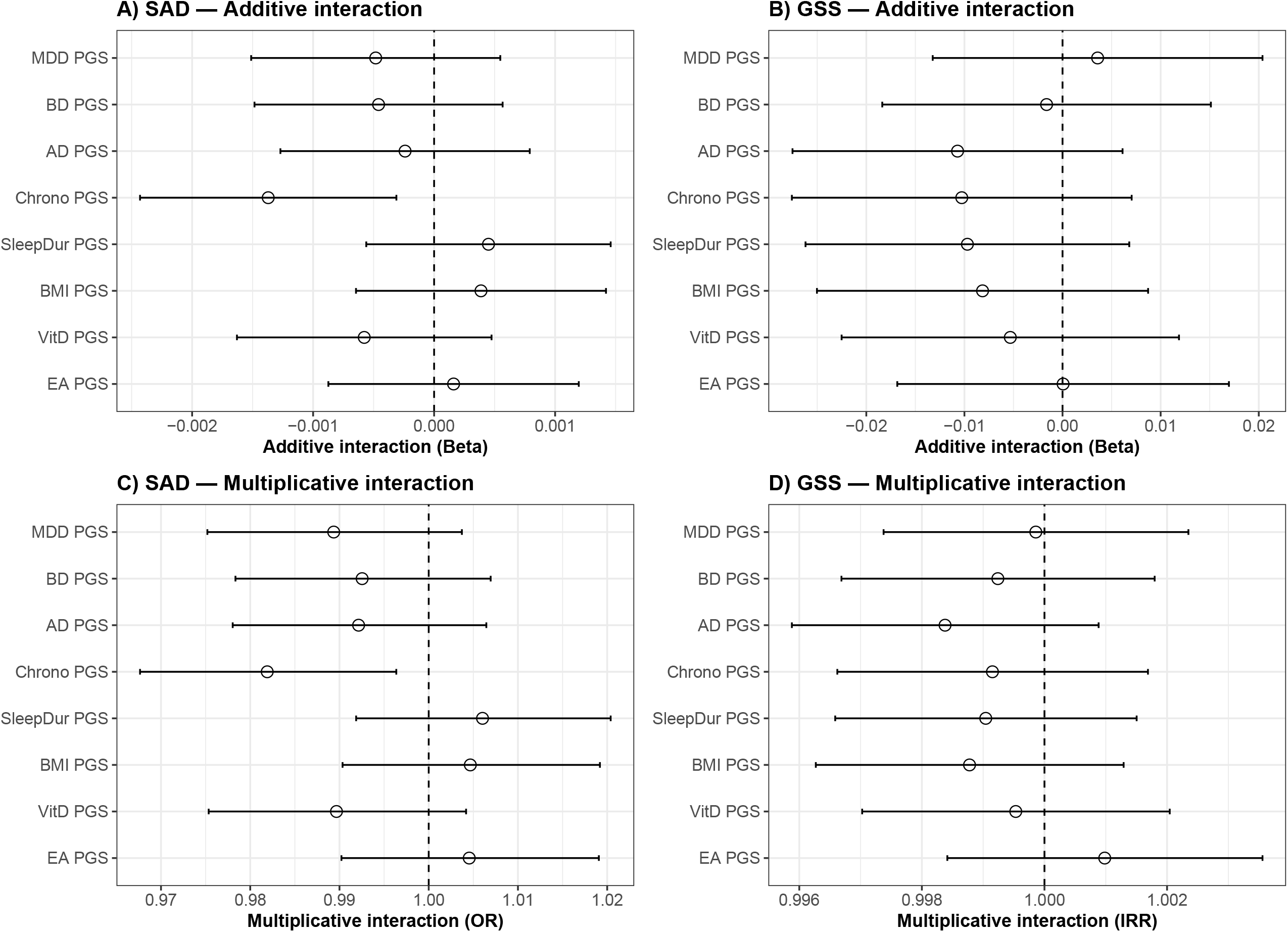
Additive and multiplicative interaction effects of polygenic scores and latitude on seasonality. Results from polygenic score-by-latitude interaction models predicting seasonal affective disorder (SAD) and general seasonality score (GSS). A) Additive interaction models predicting SAD (N = 12,460). B) Additive interaction models predicting GSS (N = 12,460). C) Multiplicative interaction models predicting SAD (N = 12,460). D) Multiplicative interaction models predicting GSS (N = 12,460). Error bars represent 95% confidence intervals. A filled in circle indicates significance after multiple testing correction (Benjamini-Hochberg False Discovery Rate P < 0.05). Abbreviations: MDD = major depressive disorder. PGS = polygenic score. BD = bipolar disorder. AD = anxiety disorders. Chrono = chronotype. SleepDur = sleep duration. BMI = body mass index. VitD = vitamin D. EA = educational attainment. OR = odds ratio. IRR = incidence rate ratio.

### Sensitivity Analyses

Controlling for MDD status (i.e., meeting CIDI short-form criteria for lifetime MDD) somewhat attenuated the already nonsignificant positive association between MDD-PGS and SAD, which remained nonsignificant across all models. The association between MDD-PGS and GSS was also slightly attenuated but remained significant in each of the MDD severity-corrected models (see Supplementary Text, Sensitivity analyses results and Table S3). Our results suggest the association between genetic risk for MDD and seasonality may in part reflect MDD severity, but that there may also be a unique relationship beyond severity.

Results for the alternative definition of SAD following the example of Kasper et al.(1), where only two months of winter and summer were considered in determining SAD case-control status (rather than three as used in our main analyses), were similar to the main model (OR=1.08[1.01-1.15], p_adjusted_=0.10) (Table S3). The association between genetic risk for MDD and SAD was considerably stronger and became significant when the ‘feel worst’ calendar item was not considered in determining SAD case-control status (OR=1.12[1.06-1.19], p_adjusted_=0.002).

## Discussion

Seasonal affective disorder (SAD) is a complex condition whose prevalence and burden are well-characterized. However, its etiology and nosological status as an MDD specifier versus a transdiagnostic disorder are the subject of active debate (7,10). In this study, we investigated the underlying etiology of SAD and dimensional seasonality by examining their association with genetic risk for a range of traits, latitude and their interaction within a large, well-characterized Australian cohort of individuals with a history of depression. Our findings show that latitude is positively associated with both SAD case-control status and the continuous dimension of seasonality, the general seasonality score (GSS). Polygenic scores for major depressive disorder (MDD), bipolar disorder (BD), anxiety disorders, vitamin D levels, and educational attainment were associated with GSS as a continuous measure of seasonality, but not with SAD case–control status. By focusing on a sample of individuals with depression, our analyses offer insight into differential vulnerability to seasonally patterned symptoms within this vulnerable group. Our multi-trait genetic and categorical-dimensional approaches provide new insights into the genetic and environmental factors shaping seasonality and its clinical manifestations.

We found that living further from the equator (higher latitude) was positively associated with higher odds of SAD and higher GSS values among individuals with seasonality symptoms. This finding aligns with theories on the effect of photoperiod on seasonal changes in behavior and mood (14). The only other study on latitude and seasonality in the southern hemisphere (Brazil, using quartile-based divisions of latitude) did not find an association between latitude and seasonality (19). It is possible that our continuous measure of latitude may capture more information, or that the variability in latitude effects on seasonality seen across Northern Hemisphere populations also applies to Southern Hemisphere countries (20). Our results indicate that latitude and photoperiod may be an essential consideration when studying seasonality.

We found significant positive associations between psychiatric polygenic scores and seasonality. These effects were slightly attenuated when examined together, particularly for MDD and anxiety disorders, suggesting part of the predictive effect of psychiatric genetic predisposition is shared. This is unsurprising given diagnostic comorbidity (35,36), though it is a novel finding in the context of seasonality. It is striking that, in a cohort of individuals with a history of depression, the genetic risk for mood and anxiety disorders still explains significant variation. One explanation is that seasonality reflects a more severe rendition of MDD and BD. However, this is contrasted by our finding that the association between the polygenic score for MDD and seasonality is partly independent of depression severity. Further, we must consider that MDD severity might not be independent of season. For example, if MDD severity and seasonality are correlated (having more symptoms or severity during, e.g., winter), then controlling for severity takes out part of the seasonality effect. An alternative explanation is that the depression genome-wide association study on which the polygenic score is based might partly reflect seasonality; up to 22% of depressive episodes have a seasonality component and so it is possible that the polygenic score captures some of this signal (10). We also observed that genetic liability for educational attainment had a protective effect for both SAD and GSS. While the individual-level interactions between genetic risk, behaviour, and illness are likely to be complex, a broad interpretation is that genetic liability for mental disorders and lower educational attainment may lead to behavioural changes that affect exposure and entrainment to the environment. In the case of genetic risk for mental disorders, it is also possible that part of the genetic contribution affects specific sensitivities to the environment (e.g., light sensitivity) which in turn produce variation in seasonality(37).

A higher genetic predisposition to increased vitamin D levels significantly decreased the probability of having no seasonality symptoms, meaning it only explained differences in whether any or no seasonality symptoms are present, but not variation in the number of seasonality symptoms. One explanation is that those who go outside more will be more exposed to environmental patterns, thus being more likely to experience any changes in behavior due to the seasons. Altogether, there may be several unmeasured behavioral and physiological (e.g., light sensitivity) factors which may interact and compete in their effect on the expression of seasonality.

We found no link between seasonality and genetic risk for sleep duration and body mass index (BMI). Up to 80% of SAD patients experience hypersomnia (38), making the absence of an effect surprising. However, the change in sleep duration with seasons may merely be a byproduct of the actual pathophysiological mechanism, meaning genetic risk for sleep duration would not be expected to play a role. Mood disorders incorporate aspects of weight and appetite in their symptomatology, but the absence of an effect could indicate that genetic risk for BMI does not explain seasonal changes in these traits. Alternatively, the relationship between BMI and depression symptoms is known to be non-linear (39), and this may cloud the signal with seasonality as well.

Overall, we observed no genetic risk by latitude interaction effects on seasonality. We did detect an interaction between genetic propensity for chronotype and latitude on SAD, but this association did not survive correction for multiple testing and should therefore be interpreted cautiously. Specifically, individuals with a genetic tendency toward morning chronotype showed a weaker association between higher latitude and increased SAD risk. One possible interpretation is that at higher latitudes, where seasonal changes in daylight are more pronounced, those with an evening chronotype may experience greater circadian disruption, potentially heightening vulnerability to SAD. This finding will need replication, and we highlight it as a hypothesis-generating result.

We found clear differences between SAD and GSS in associations with polygenic scores, in that polygenic score-seasonality associations were only evident for GSS. Dimensional measures such as the GSS may better capture underlying genetic influences than binary classifications, which impose diagnostic cut-offs that do not necessarily reflect the continuous nature of mood and seasonality variation. Alternatively, the SAD criteria involving severity and ‘worst month’ calendar item have been criticized and may warrant additional consideration (40). This was also reflected in the comparison of SAD definitions, where dropping the ‘worst month’ criterion revealed a positive genetic association with MDD PGS after multiple testing correction. These findings align with recent discussions on SAD assessment and seasonality measurement (e.g., (41,42)), which highlight that the SPAQ is a useful dimensional instrument but not a strict diagnostic classifier and that its underlying factor structure may warrant further investigation.

Several limitations are present for this study. Analyses were limited to individuals of European ancestry, those with severe and recurrent depression and those of higher education (30), which may limit the study’s generalizability. Further, the relative power of each polygenic score is influenced by the sample size of the GWAS and genetic architecture of the phenotype, which may affect the difference in associations (43). While several polygenic scores were significantly associated, effects were generally small, which may be due to a combination of cross-phenotype analyses and the somewhat narrow definition of SAD and seasonality. Moreover, latitude is only one aspect of geographical location that may influence seasonality. A fuller picture would require incorporating climatic variation, direct photoperiod measures, and individual differences in light-exposure behaviors, which latitude alone cannot capture. Finer-grained regional factors such as microclimate or degree of urbanicity may also influence how seasonality is experienced, and should be explored in future studies. Exploring seasonality among individuals with a history of depression gives insight into depression heterogeneity but may also reduce generalizability to the general population if we consider SAD to be a transdiagnostic syndrome. It would be interesting to repeat these analyses in a general population sample. Larger sample sizes will also benefit the study of interaction analyses, and further studies are needed to replicate these results.

In the first study of its kind, we identified significant associations between latitude, genetic risk for MDD, BD, anxiety disorders, educational attainment and vitamin D levels with seasonality. These findings advance our understanding of the etiology of SAD as a multifactorial trait by emphasizing the contribution of psychiatric genetic predisposition and environmental factors. The associations observed with the GSS underscore the value of examining seasonality as a continuous dimension, offering potential insights beyond categorical classifications.

## Data Availability

All data produced in the present study are available upon reasonable request to the authors

## Acknowledgments

We are indebted to the effort and time that AGDS participants gracefully gave to contribute to this study. We would like to thank the research participants and employees of 23andMe, Inc. for making this work possible. The genome-wide summary statistics for the Jones et al. (2019) and O’Connell et al. (2025) analysis of 23andMe, Inc., data were obtained under a data transfer agreement with the QIMR Berghofer Medical Research Institute. We also thank those involved in conception, implementation, beta testing, media campaign, and data cleaning. Special thanks go to Naomi Wray for advice and contributions to the study design; Katherine Kirk for project management; Richard Parker for project coordination; Lorelle Nunn, Rebekah Cicero, Mary Ferguson, Lucy Winkler, and Natalie Garden for data and sample collection; Simone Cross for data collection and project management; Lenore Sullivan for participant contact; Natalia Zmicerevska, Alissa Nichles, and Candace Brennan for recruitment support; Jonathan Davies, Luke Lowrey, and Valeriano Antonini for IT support; Vera Morgan and Ken Kirkby for media campaign assistance; Scott Gordon for carrying out the imputation and quality control of the genotype data; and Penelope Lind for contribution to the management of the AGDS database. The Australian Genetics of Depression Study was primarily funded by grant 1086683 from the National Health and Medical Research Council (NHMRC) of Australia. This work was supported by NHMRC Investigator Grants to JJC (2008196); BLM (2017176); SEM (1172917); NGM (1172990); and IBH (2016346). Role of the Funder/Sponsor: The funding organizations had no role in the design and conduct of the study; collection, management, analysis, and interpretation of the data; preparation, review, or approval of the manuscript; and decision to submit the manuscript for publication.

## Author contributions

*Concept and design:* Huider, Crouse, Thomas, Mitchell.

*Acquisition, analysis, or interpretation of data:* Huider, Medland, Martin, Hickie, Crouse, Thomas, Mitchell.

*Intellectual content:* Huider, Medland, Martin, Hickie, Crouse, Thomas, Mitchell.

*Statistical analysis:* Huider, Thomas, Mitchell.

*Obtained funding:* Medland, Hickie, Martin, Mitchell.

*Administrative, technical, or material support:* Huider, Thomas, Mitchell.

*Supervision:* Thomas, Mitchell.

## Data Availability

Data are available upon reasonable request via the corresponding authors.

