## Supplementary Materials for "Genetic and Environmental Predictors of Seasonality and Seasonal Affective Disorder in Individuals with Depression"

**Supplementary Text.** Psychometric information for the SPAQ.

**Figure S1**. Decision tree for deriving seasonal affective disorder (SAD) case-control status.

**Supplementary Text.** Genotype information and polygenic score generation**.**

**Supplementary Text.** Model selection for general seasonality score.

**Figure S2**. Distribution of general seasonality score (N = 12,613).

**Supplementary Text.** Sensitivity analyses methods.

**Figure S3.** Flowchart of study sample selection.

**Figure S4**. Prevalence of SPAQ-item 'feeling worst' by month.

**Table S1.** Univariate and multivariate polygenic score and latitude prediction of seasonality.

**Table S2.** Results from multivariate prediction of seasonality with polygenic scores and latitude, and their multiplicative and additive interaction.

**Supplementary Text.** Sensitivity analyses results.

**Table S3.** Results from MDD polygenic score prediction of seasonality corrected for MDD severity and alternate SAD criteria.

**Supplementary References**

This supplemental material has been provided by the authors to give readers additional information about their work.

**Supplementary Text.**

**Psychometric information for the SPAQ.**

The Seasonal Pattern Assessment Questionnaire (SPAQ) is a self-assessment instrument that was developed by Rosenthal et al. (1) to measure seasonal changes in mood and behavior. It is the most frequently used questionnaire for seasonal affective disorder (SAD) and has been translated to different languages (2–4), with the benefit that it can be used in both general and clinical populations thanks to phrasing and non assumption of mood disruption or disease (5). The SPAQ was found to have a one-factor structure (6) or two-factor structure (4,7), where the latter identifies a psychological factor component for the items mood, sleep length, energy and social activity, and a food factor for weight and appetite.

Ambar Akkaoui & Geoffroy (2025) recently completed a systematic review of the psychometric properties of the SPAQ (8). In the 28 studies they compared, 16 focused on the SPAQ. Sensitivity estimates in previous studies ranged between 40% and 94% for sensitivity, depending on the cut off criteria for determining SAD (7,9,10), Specificity estimates ranged between 46% and 94%. Positive predictive values ranged between 45% and 71%, negative predictive values between 46% and 83%, and the SPAQ has demonstrated moderate to high test–retest reliability (TRRs ranging from 0.51 to 0.87), supporting stability of the measure. Finally, internal consistency (Cronbach alpha) estimates ranged between 0.74 and 0.86, and inter-judge kappa values between 0.44 and 0.79 (8).

**Figure S1. Decision tree for deriving seasonal affective disorder (SAD) case-control status.**

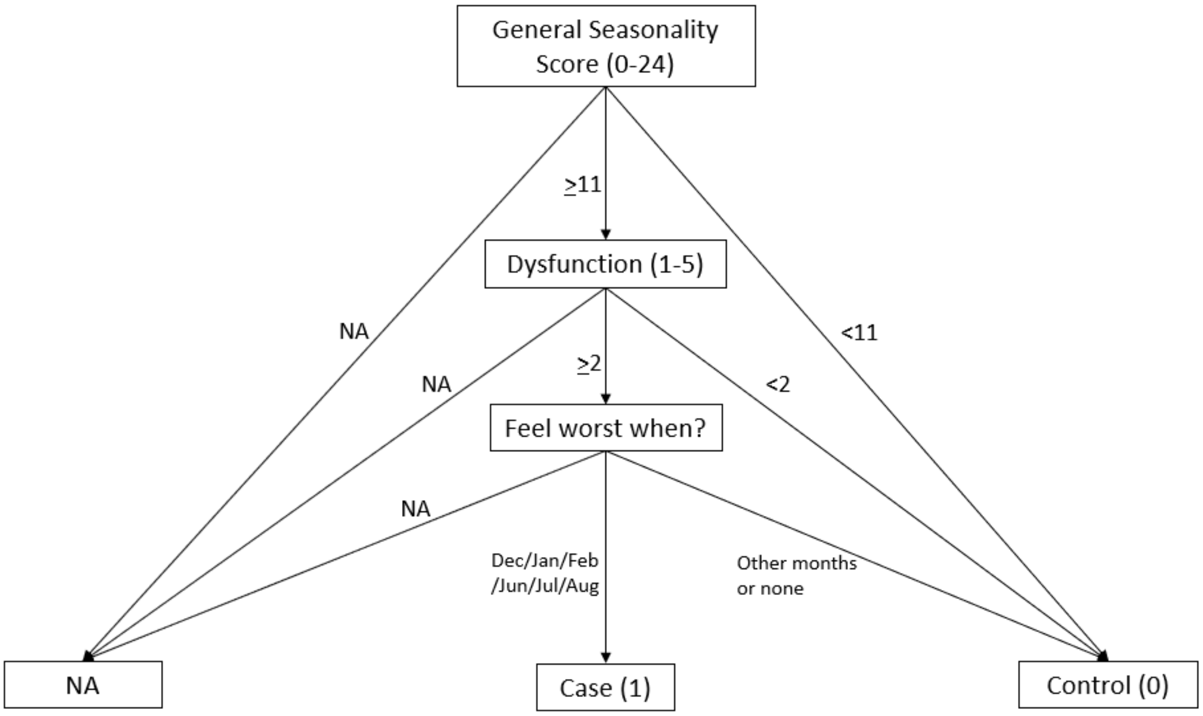


Measures derived from the Seasonal Pattern Assessment Questionnaire. Dysfunction levels 1-5 correspond to mild, moderate, marked, severe, and disabling, respectively.

**Supplementary Text.**

**Genotype information and polygenic score generation.**

Genotyping was performed using the Illumina Global Screening Array v2. Low-quality genotype samples were removed; chromosomal sex was inferred from X-chromosome heterozygosity, and individuals with discrepancies between inferred and reported sex were excluded. The remaining samples were merged with those from the 1000 Genomes Project for principal component calculation using 'smartpca' (build 16000) in a set of unlinked single nucleotide polymorphisms (SNPs). Pre-imputation quality control (QC) was conducted using GenomeStudio v2.0 and PLINK 1.9 (11), and involved the exclusion of SNPs with a minor allele frequency (MAF) < 0.01, a call rate < 95%, and a Hardy-Weinberg equilibrium (*P* < 1.00E-6), and samples with a non-European ancestry defined as >4 standard deviations from the ancestry principal component (PC) centroid for PC1 and PC2. Imputation was then performed using the Haplotype Reference Consortium 1.1 reference panel (12).

Polygenic scores (PGSs) were generated using SBayesR. SBayesR is a Bayesian approach that assumes that GWAS single nucleotide polymorphism (SNP) effects are sampled from a mixture of four zero-mean normal distributions with varying variances, with many GWAS SNPs assumed to have effect sizes of zero (13). The default SBayesR settings in GCTB were applied, utilizing a mixture of four normal distributions with $\pi$ values of 0.95, 0.03, 0.01, and 0.01, and variance scaling factors of 0.0, 0.01, 0.1, and 1 for the respective components. Variants within the Major Histocompatibility Complex were excluded from the analysis. The chain length was set to 25,000 iterations, with a burn-in of 5,000, and an initial heritability estimate of 0.5 was used for all traits. For the LD reference, we employed the same sparse LD matrix as in Lloyd-Jones et al. (13), constructed from HapMap3 SNPs of 50,000 randomly selected, unrelated individuals from the UKBiobank. The resulting posterior SNP effects were used as input in the PLINK 1.9 score function to generate PGS (11).

**Supplementary Text.**

**Model selection for general seasonality score.**

We used the “performance”(14), “DHARMa”(15), “lmtest”(16), and “VGAM”(17) R packages to assess model fit and assumptions for the general seasonality score (GSS) regression models, using major depressive disorder (MDD) polygenic score (PGS) as a predictor and sex, age, and 10 principal components as covariates. The skewed count distribution of GSS led us to consider Poisson regression, negative binomial regression, and zero-inflated negative binomial regression (mixed and hurdle models). Starting with the Poisson regression, we found the results to be heavily overdispersed (dispersion = 3.2207, P-value = 2.2E-16). Next, we tested negative binomial regression. The negative binomial model had a lower AIC (ΔAIC = -14,361.1), and a significantly better fit than the Poisson model (Likelihood ratio test $Chi^2$ = 14366, P-value = 2.2E-16). However, the negative binomial model was mildly but significantly underdispersed (dispersion: 0.8119, P-value = 2.2E-16) and the model performed poorly at capturing the excess of zeros in the GSS distribution. Next, we fit a zero-inflated negative binomial model to the data, finding a dispersion estimate of 0.961 ($Chi^2$ P-value = 0.999). Model fit between the negative binomial and zero-inflated negative binomial model was compared using the glmmTMB framework and evaluated them with DHARMa simulation-based tests. The negative binomial model showed evidence of unmodelled zero inflation (zero-inflation test ratioObsSIm = 2.02, P-value < 2.2E-16) and underdispersion (dispersion = 0.81, P-value < 2.2E-16). In contrast, the zero-inflated negative binomial model showed no evidence of excess zeros (ratioObsSIm = 1.01, P-value = 0.784), near-ideal dispersion (dispersion = 0.95), and substantially better fit as indexed by information criteria ($AIC_{NB}$ = 72,606.6; $AIC_{ZINB}$ = 71,551.3; ΔAIC = -1.055). Finally, we compared model performance for a two-component mixture zero-inflated negative binomial model and a two-step (hurdle) zero-inflated negative binomial model. The fit of the two models did not differ significantly (Vuong AIC-corrected z-statistic of 0.469, P-value = 0.320), and the lower AIC of the mixture zero-inflated negative binomial model (ΔAIC = -0.19) led us to choose this model.

The mixture zero-inflated negative binomial regression model has two components; 1) a count model testing the association between the predictor and the non-zero GSS values, and 2) a zero-inflated model that tests the association between a predictor and the probability of having no seasonality symptoms (GSS = 0).

**Figure S2. Distribution of general seasonality score (N = 12,613).**

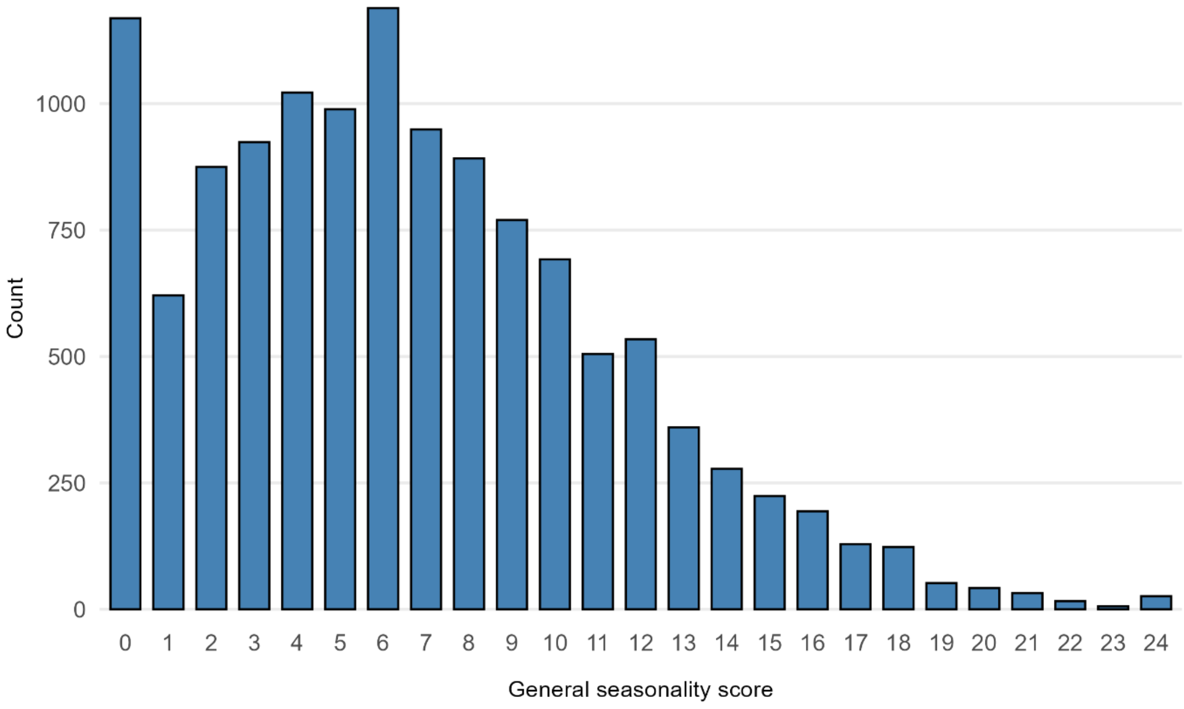

**Supplementary Text.**

**Sensitivity analyses methods.**

*MDD Severity*

To assess whether the association between seasonality and the major depressive disorder (MDD) polygenic score (PGS) depended on MDD severity, we considered three variables to serve as proxy for MDD severity. The first was the number of depressive episodes, defined as "How many periods have you had in your life where you felt depressed or lost interest in things every day or nearly every day for at least two weeks?" (1 – 12 episodes, or more than 12). The second severity measure was the number of depressive symptoms, counting the presence of MDD core and accessory symptoms (0 – 9). The third severity measure was the duration of the worst depressive episode, measured in an open question as the number of weeks. Only responses where the number of weeks did not exceed the age at measurement were included. We then predicted SAD in logistic regression with MDD PGS and number of depressive episodes (and sex, age, and 10 principal components). We repeated this approach for the other severity measures, number of depressive symptoms and duration of the worst depressive episode, and for GSS in linear regression.

*SAD Definition*

To assess whether our deviation from Kasper et al.'s(18) definition of seasonal affective disorder (SAD) affected MDD PGS prediction, we analyzed an alternative definition of SAD where cases were limited to those who felt worst during January or February, and July or August (as opposed to all three months of meteorological summer and winter). Second, to assess the relevance of the 'feel worst' calendar criterion in SAD case-control status, we created an alternative definition of SAD where this criterion was dropped and tested the association with MDD polygenic score. We ran logistic regression models predicting these alternative definitions of SAD with MDD PGS (and sex, age, and 10 principial components).

**Figure S3. Flowchart of study sample selection.**

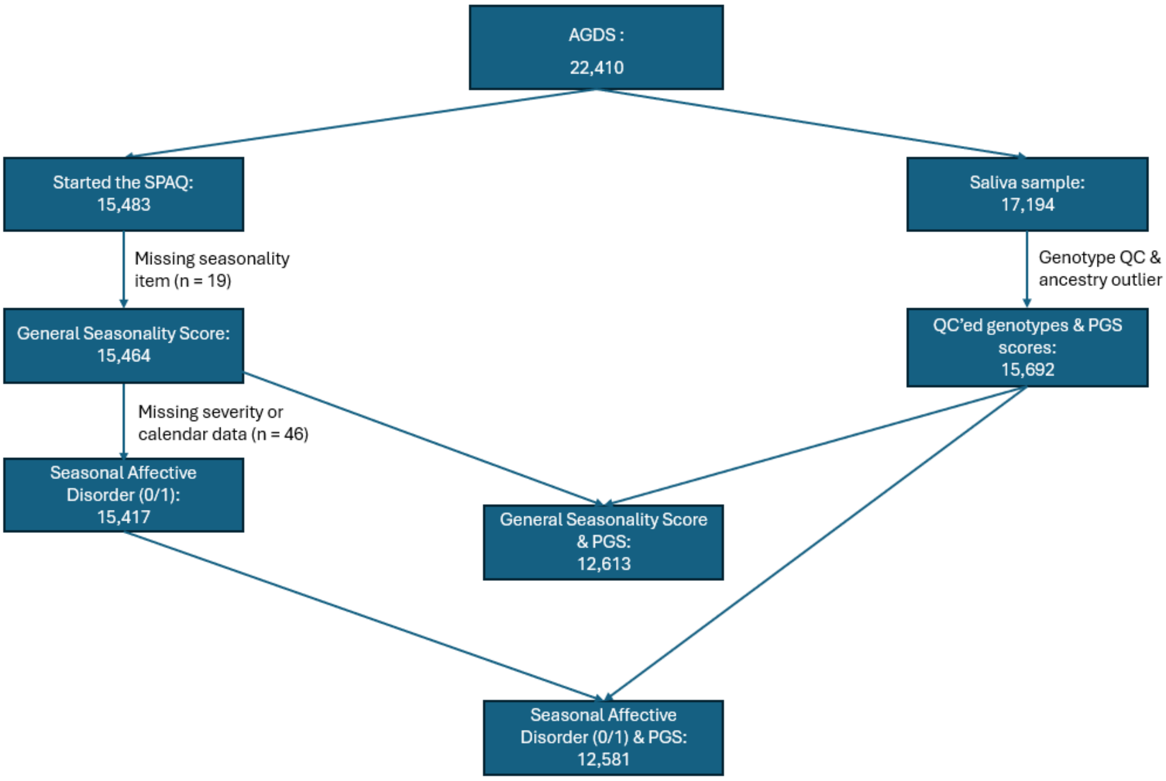


Abbreviations: AGDS = Australian Genetics of Depression Study. SPAQ = Seasonal Pattern Assessment Questionnaire. PGS = polygenic score. QC = quality control.

**Figure S4. Prevalence of individuals who indicate 'feeling worst' during a particular month. Data were double-plotted to facilitate visual interpretation.**

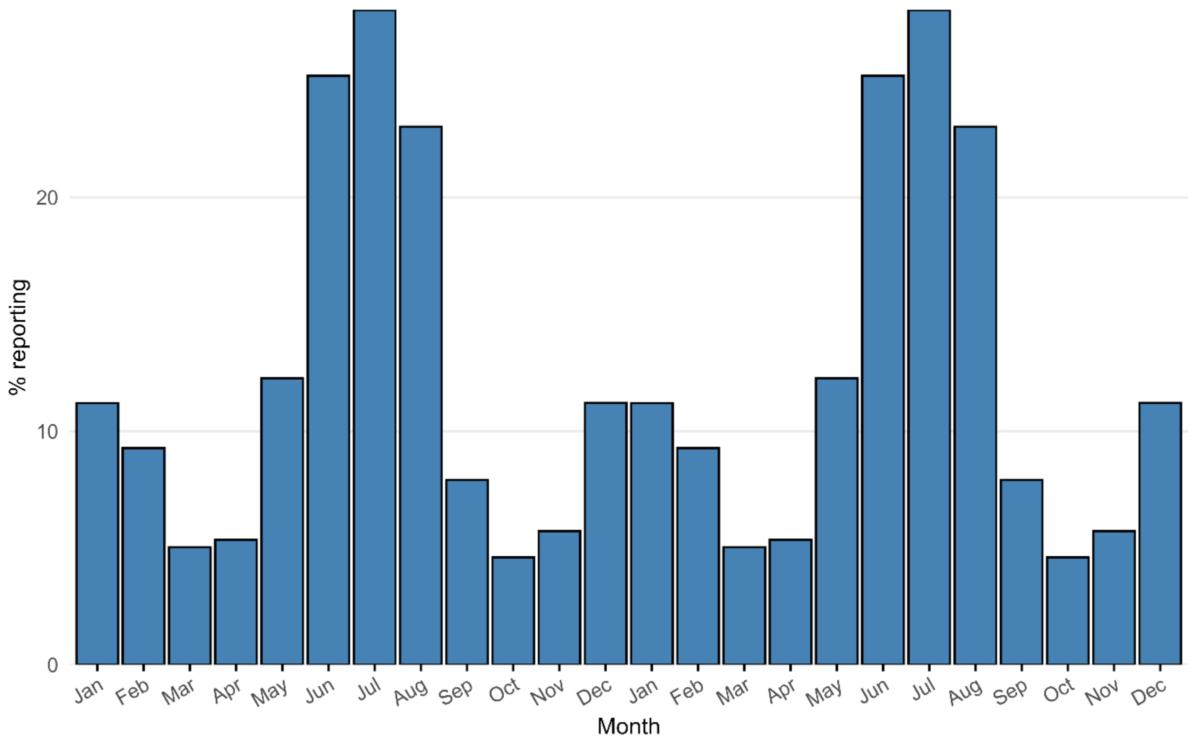

**Table S1: Single and multiple polygenic score and latitude prediction of seasonality.**

| Outcome | Model | Predictor | Effect | SE | CI | P | P_adjusted |
|---|---|---|---|---|---|---|---|
| SAD | Single-PGS | MDD PGS | 1.0786 | 0.0321 | 1.0130 – 1.1486 | **0.0182** | 0.124 |
| | Single-PGS | BD PGS | 0.9948 | 0.0318 | 0.9346 – 1.0589 | 0.87 | 0.87 |
| | Single-PGS | AD PGS | 1.0678 | 0.0319 | 1.0031 – 1.1367 | **0.0397** | 0.193 |
| | Single-PGS | Chrono PGS | 0.942 | 0.0321 | 0.8845 – 1.0033 | 0.0632 | 0.269 |
| | Single-PGS | SleepDur PGS | 1.0473 | 0.0319 | 0.9839 – 1.1149 | 0.147 | 0.337 |
| | Single-PGS | BMI PGS | 1.0237 | 0.0319 | 0.9616 – 1.0898 | 0.463 | 0.578 |
| | Single-PGS | VitD PGS | 1.0358 | 0.0322 | 0.9725 – 1.1032 | 0.274 | 0.479 |
| | Single-PGS | EA PGS | 0.9549 | 0.032 | 0.8969 – 1.0166 | 0.149 | 0.337 |
| | Single predictor | Latitude | 1.0484 | 0.0073 | 1.0335 – 1.0635 | **0.000000000861** | **0.00000000293** |
| | Multiple-PGS | MDD PGS | 1.0648 | 0.0377 | 0.9890 – 1.1464 | 0.0959 | 0.296 |
| | Multiple-PGS | BD PGS | 0.9763 | 0.0328 | 0.9155 – 1.0412 | 0.465 | 0.578 |
| | Multiple-PGS | AD PGS | 1.0339 | 0.0378 | 0.9602 – 1.1133 | 0.377 | 0.543 |
| | Multiple-PGS | Chrono PGS | 0.9429 | 0.0323 | 0.8850 – 1.0046 | 0.0692 | 0.243 |
| | Multiple-PGS | SleepDur PGS | 1.0542 | 0.0322 | 0.9898 – 1.1229 | 0.101 | 0.312 |
| | Multiple-PGS | BMI PGS | 1.0293 | 0.0321 | 0.9665 – 1.0962 | 0.369 | 0.543 |
| | Multiple-PGS | VitD PGS | 1.0444 | 0.0323 | 0.9803 – 1.1128 | 0.179 | 0.38 |
| | Multiple-PGS | EA PGS | 0.9539 | 0.0331 | 0.8940 – 1.0177 | 0.153 | 0.345 |
| GSS | Single-PGS (NB) | MDD PGS | 1.0279 | 0.006 | 1.0159 – 1.0400 | **0.00000429** | **0.0000362** |
| | Single-PGS (ZI) | MDD PGS | 1.0751 | 0.0377 | 0.9986 – 1.1575 | 0.0544 | 1 |
| | Single-PGS (NB) | BD PGS | 1.0222 | 0.006 | 1.0102 – 1.0343 | **0.000269** | **0.00102** |
| | Single-PGS (ZI) | BD PGS | 1.0614 | 0.0378 | 0.9855 – 1.1431 | 0.115 | 1 |
| | Single-PGS (NB) | AD PGS | 1.0269 | 0.006 | 1.0149 – 1.0391 | **0.0000101** | **0.0000493** |

| | | | | | | |
|---|---|---|---|---|---|---|
| Single-PGS (ZI) | AD PGS | 1.0566 | 0.0382 | 0.9804 – 1.1388 | 0.149 | 1 |
| Single-PGS (NB) | Chrono PGS | 0.9943 | 0.006 | 0.9827 – 1.0059 | 0.334 | 0.687 |
| Single-PGS (ZI) | Chrono PGS | 0.9809 | 0.0377 | 0.9110 – 1.0561 | 0.609 | 1 |
| Single-PGS (NB) | SleepDur PGS | 1.0006 | 0.006 | 0.9890 – 1.0124 | 0.919 | 0.976 |
| Single-PGS (ZI) | SleepDur PGS | 1.0128 | 0.0377 | 0.9406 – 1.0906 | 0.735 | 1 |
| Single-PGS (NB) | BMI PGS | 0.9986 | 0.006 | 0.9869 – 1.0103 | 0.809 | 0.976 |
| Single-PGS (ZI) | BMI PGS | 1.0236 | 0.0379 | 0.9503 – 1.1026 | 0.538 | 1 |
| Single-PGS (NB) | VitD PGS | 0.9986 | 0.006 | 0.9870 – 1.0104 | 0.817 | 0.976 |
| Single-PGS (ZI) | VitD PGS | 0.8885 | 0.0371 | 0.8262 – 0.9554 | **0.00142** | **0.0484** |
| Single-PGS (NB) | EA PGS | 0.9731 | 0.006 | 0.9618 – 0.9846 | **0.00000532** | **0.0000362** |
| Single-PGS (ZI) | EA PGS | 0.8453 | 0.0384 | 0.7841 – 0.9114 | **0.0000119** | **0.000406** |
| Single predictor (NB) | Latitude | 1.0116 | 0.0013 | 1.0090 – 1.0141 | **1.62e-19** | **5.51e-18** |
| Single predictor (ZI) | Latitude | 0.9495 | 0.007 | 0.9365 – 0.9627 | **0.0000000000018** | **0.00000000000598** |
| Multiple-PGS (NB) | MDD PGS | 1.0158 | 0.007 | 1.0020 – 1.0297 | **0.0243** | 0.0827 |
| Multiple-PGS (NB) | BD PGS | 1.0159 | 0.0062 | 1.0037 – 1.0282 | **0.0103** | **0.0351** |
| Multiple-PGS (NB) | AD PGS | 1.011 | 0.007 | 0.9972 – 1.0250 | 0.12 | 0.407 |
| Multiple-PGS (NB) | Chrono PGS | 0.9938 | 0.0059 | 0.9823 – 1.0055 | 0.299 | 0.687 |
| Multiple-PGS (NB) | SleepDur PGS | 1.0036 | 0.006 | 0.9920 – 1.0154 | 0.542 | 0.828 |
| Multiple-PGS (NB) | BMI PGS | 1.0012 | 0.0059 | 0.9896 – 1.0130 | 0.836 | 0.976 |
| Multiple-PGS (NB) | VitD PGS | 1.001 | 0.0059 | 0.9894 – 1.0127 | 0.871 | 0.976 |
| Multiple-PGS (NB) | EA PGS | 0.9754 | 0.0061 | 0.9638 – 0.9872 | **0.0000508** | **0.000216** |

In the single-PGS models, each model contained latitude or the polygenic score for one trait (nine models for each outcome). In the multiple-PGS models, all eight polygenic scores were included in the same model. *P*-adj = Multiple testing-adjusted P-value (Benjamini-Hochberg False Discovery Rate $P < 0.05$). SAD = seasonal affective disorder. GSS = General seasonality score. MDD = major depressive disorder. PGS = polygenic score. BD = bipolar disorder. AD = anxiety disorders. BMI =

body-mass index. EA = educational attainment. PGS = polygenic score. NB = count part of the zero-inflated negative binomial model. ZI = zero-inflated part of the negative binomial model. Bold indicates nominal significance (P < 0.05) or significance (FDR P < 0.05).

**Table S2: Results from multivariate prediction of seasonality with polygenic scores and latitude, and their multiplicative and additive interaction.**

| Outcome | Model | Predictor | Effect | SE | CI | P | P_adjusted |
|---|---|---|---|---|---|---|---|
| SAD | Latitude-adjusted | MDD PGS | 1.082 | 0.0321 | 1.0160 – 1.1522 | **0.0141** | 0.122 |
| | Latitude-adjusted | BD PGS | 0.9948 | 0.0319 | 0.9345 – 1.0589 | 0.869 | 0.87 |
| | Latitude-adjusted | AD PGS | 1.0684 | 0.0319 | 1.0036 – 1.1374 | **0.0382** | 0.193 |
| | Latitude-adjusted | Chrono PGS | 0.9436 | 0.0322 | 0.8859 – 1.0051 | 0.0714 | 0.27 |
| | Latitude-adjusted | SleepDur PGS | 1.0477 | 0.032 | 0.9840 – 1.1156 | 0.145 | 0.337 |
| | Latitude-adjusted | BMI PGS | 1.0231 | 0.032 | 0.9609 – 1.0893 | 0.476 | 0.578 |
| | Latitude-adjusted | VitD PGS | 1.0412 | 0.0322 | 0.9775 – 1.1090 | 0.21 | 0.421 |
| | Latitude-adjusted | EA PGS | 0.9458 | 0.032 | 0.8883 – 1.0071 | 0.0821 | 0.279 |
| | Additive interaction | MDD PGS | -0.0005 | 0.0005 | -0.0015 – 0.0005 | 0.358 | 0.543 |
| | Additive interaction | BD PGS | -0.0005 | 0.0005 | -0.0015 – 0.0006 | 0.38 | 0.543 |
| | Additive interaction | AD PGS | -0.0002 | 0.0005 | -0.0013 – 0.0008 | 0.648 | 0.711 |
| | Additive interaction | Chrono PGS | -0.0014 | 0.0005 | -0.0024 – -0.0003 | **0.0112** | 0.122 |
| | Additive interaction | SleepDur PGS | 0.0004 | 0.0005 | -0.0006 – 0.0015 | 0.383 | 0.543 |
| | Additive interaction | BMI PGS | 0.0004 | 0.0005 | -0.0006 – 0.0014 | 0.462 | 0.578 |
| | Additive interaction | VitD PGS | -0.0006 | 0.0005 | -0.0016 – 0.0005 | 0.282 | 0.479 |
| | Additive interaction | EA PGS | 0.0002 | 0.0005 | -0.0009 – 0.0012 | 0.761 | 0.808 |
| | Multiplicative interaction | MDD PGS | 0.9894 | 0.0074 | 0.9752 – 1.0037 | 0.146 | 0.337 |
| | Multiplicative interaction | BD PGS | 0.9925 | 0.0074 | 0.9783 – 1.0069 | 0.308 | 0.499 |
| | Multiplicative interaction | AD PGS | 0.9921 | 0.0073 | 0.9780 – 1.0065 | 0.28 | 0.479 |
| | Multiplicative interaction | Chrono PGS | 0.9819 | 0.0075 | 0.9677 – 0.9964 | **0.0144** | 0.122 |
| | Multiplicative interaction | SleepDur PGS | 1.006 | 0.0072 | 0.9919 – 1.0204 | 0.406 | 0.552 |
| | Multiplicative interaction | BMI PGS | 1.0047 | 0.0073 | 0.9903 – 1.0192 | 0.525 | 0.608 |

| | | | | | | | |
|---|---|---|---|---|---|---|---|
| | Multiplicative interaction | VitD PGS | 0.9897 | 0.0074 | 0.9753 – 1.0042 | 0.162 | 0.345 |
| | Multiplicative interaction | EA PGS | 1.0045 | 0.0073 | 0.9902 – 1.0191 | 0.536 | 0.608 |
| GSS | Latitude-adjusted (NB) | MDD PGS | 1.0285 | 0.006 | 1.0166 – 1.0406 | **0.00000237** | **0.0000269** |
| | Latitude-adjusted (ZI) | MDD PGS | 1.073 | 0.0378 | 0.9965 – 1.1555 | 0.062 | 1 |
| | Latitude-adjusted (NB) | BD PGS | 1.0223 | 0.006 | 1.0104 – 1.0344 | **0.000232** | **0.000986** |
| | Latitude-adjusted (ZI) | BD PGS | 1.0617 | 0.0379 | 0.9857 – 1.1436 | 0.114 | 1 |
| | Latitude-adjusted (NB) | AD PGS | 1.0274 | 0.006 | 1.0154 – 1.0396 | **0.00000646** | **0.0000366** |
| | Latitude-adjusted (ZI) | AD PGS | 1.0566 | 0.0383 | 0.9801 – 1.1391 | 0.151 | 1 |
| | Latitude-adjusted (NB) | Chrono PGS | 0.9949 | 0.0059 | 0.9834 – 1.0065 | 0.388 | 0.733 |
| | Latitude-adjusted (ZI) | Chrono PGS | 0.9776 | 0.0379 | 0.9077 – 1.0529 | 0.55 | 1 |
| | Latitude-adjusted (NB) | SleepDur PGS | 1.0009 | 0.0059 | 0.9893 – 1.0127 | 0.876 | 0.976 |
| | Latitude-adjusted (ZI) | SleepDur PGS | 1.0096 | 0.0377 | 0.9377 – 1.0869 | 0.8 | 1 |
| | Latitude-adjusted (NB) | BMI PGS | 0.9985 | 0.0059 | 0.9869 – 1.0102 | 0.801 | 0.976 |
| | Latitude-adjusted (ZI) | BMI PGS | 1.0261 | 0.038 | 0.9525 – 1.1054 | 0.497 | 1 |
| | Latitude-adjusted (NB) | VitD PGS | 1.0001 | 0.0059 | 0.9885 – 1.0118 | 0.993 | 0.995 |
| | Latitude-adjusted (ZI) | VitD PGS | 0.8817 | 0.0372 | 0.8197 – 0.9485 | **0.000728** | **0.0248** |
| | Latitude-adjusted (NB) | EA PGS | 0.9708 | 0.006 | 0.9595 – 0.9822 | **0.00000065** | **0.0000111** |
| | Latitude-adjusted (ZI) | EA PGS | 0.8547 | 0.0385 | 0.7926 – 0.9218 | **0.000046** | **0.00157** |
| | Additive interaction | MDD PGS | 0.0036 | 0.0086 | -0.0132 – 0.0204 | 0.677 | 0.959 |
| | Additive interaction | BD PGS | -0.0016 | 0.0085 | -0.0184 – 0.0151 | 0.849 | 0.976 |
| | Additive interaction | AD PGS | -0.011 | 0.0086 | -0.0275 – 0.0061 | 0.212 | 0.602 |
| | Additive interaction | Chrono PGS | -0.010 | 0.0088 | -0.0276 – 0.0070 | 0.245 | 0.606 |
| | Additive interaction | SleepDur PGS | -0.010 | 0.0084 | -0.0262 – 0.0068 | 0.249 | 0.606 |

| | | | | | | |
|---|---|---|---|---|---|---|
| Additive interaction | BMI PGS | -0.008 | 0.0086 | -0.0250 – 0.0087 | 0.344 | 0.687 |
| Additive interaction | VitD PGS | -0.005 | 0.0088 | -0.0225 – 0.0119 | 0.544 | 0.828 |
| Additive interaction | EA PGS | 0.00001 | 0.0086 | -0.0168 – 0.0170 | 0.995 | 0.995 |
| Multiplicative interaction (NB) | MDD PGS | 0.9999 | 0.0013 | 0.9974 – 1.0023 | 0.911 | 0.976 |
| Multiplicative interaction (ZI) | MDD PGS | 0.9969 | 0.0071 | 0.9832 – 1.0108 | 0.661 | 1 |
| Multiplicative interaction (NB) | BD PGS | 0.9992 | 0.0013 | 0.9967 – 1.0018 | 0.56 | 0.828 |
| Multiplicative interaction (ZI) | BD PGS | 1.0012 | 0.007 | 0.9875 – 1.0151 | 0.863 | 1 |
| Multiplicative interaction (NB) | AD PGS | 0.9984 | 0.0013 | 0.9959 – 1.0009 | 0.204 | 0.602 |
| Multiplicative interaction (ZI) | AD PGS | 1.0063 | 0.0072 | 0.9923 – 1.0205 | 0.382 | 1 |
| Multiplicative interaction (NB) | Chrono PGS | 0.9992 | 0.0013 | 0.9966 – 1.0017 | 0.512 | 0.828 |
| Multiplicative interaction (ZI) | Chrono PGS | 1.0063 | 0.0073 | 0.9920 – 1.0207 | 0.391 | 1 |
| Multiplicative interaction (NB) | SleepDur PGS | 0.999 | 0.0013 | 0.9966 – 1.0015 | 0.445 | 0.771 |
| Multiplicative interaction (ZI) | SleepDur PGS | 1.0083 | 0.007 | 0.9946 – 1.0222 | 0.235 | 1 |
| Multiplicative interaction (NB) | BMI PGS | 0.9988 | 0.0013 | 0.9963 – 1.0013 | 0.34 | 0.687 |
| Multiplicative interaction (ZI) | BMI PGS | 1.0022 | 0.0071 | 0.9884 – 1.0161 | 0.757 | 1 |
| Multiplicative interaction (NB) | VitD PGS | 0.9995 | 0.0013 | 0.9970 – 1.0020 | 0.715 | 0.972 |

| | | | | | | | |
|---|---|---|---|---|---|---|---|
| | Multiplicative interaction (ZI) | VitD PGS | 1.0033 | 0.0072 | 0.9893 – 1.0175 | 0.646 | 1 |
| | Multiplicative interaction (NB) | EA PGS | 1.001 | 0.0013 | 0.9984 – 1.0036 | 0.453 | 0.771 |
| | Multiplicative interaction (ZI) | EA PGS | 1.0018 | 0.0076 | 0.9870 – 1.0168 | 0.812 | 1 |

Each model included latitude and the polygenic score for one trait as predictors (eight models for each outcome). *P*-adj = Multiple testing-adjusted P-value (Benjamini-Hochberg False Discovery Rate *P* < 0.05). SAD = seasonal affective disorder. MDD = major depressive disorder. PGS = polygenic score. BD = bipolar disorder. AD = anxiety disorders. BMI = body-mass index. EA = educational attainment. NB = count part of the zero-inflated negative binomial model. ZI = zero-inflated part of the negative binomial model. Bold indicates nominal significance (P < 0.05) or significance (FDR P < 0.05).

**Supplementary Text.**

**Sensitivity analyses results**

To explore a possible role of severity in the association between MDD PGS and seasonality, we ran three separate MDD PGS prediction models of GSS and SAD controlling for number of depressive episodes (N = 11,265), duration of longest episode (N = 10,961) and number of depressive symptoms (N = 11,085). The number of depressive episodes and number of depressive symptoms were significantly positively associated with both GSS and SAD, whereas duration of longest episode had no significant association (eTable 3). The association between MDD PGS and GSS remained significant in each of the MDD severity-corrected models. The addition of severity to the SAD model somewhat attenuated the already nonsignificant MDD PGS estimate, which remained nonsignificant across all models. The combination of these results suggests the association between MDD PGS and seasonality may partially reflect MDD severity but that they also have a unique relation that goes beyond severity.

**Table S3: Results from MDD polygenic score prediction of seasonality corrected for MDD severity and alternate SAD criteria.**

| Outcome | Model | Predictor | Effect | SE | CI | P | P_adjusted |
|---|---|---|---|---|---|---|---|
| SAD | Longest episode-adjusted | MDD PGS | 1.0666 | 0.0333 | 0.9991 – 1.1386 | 0.0531 | 0.226 |
| | Number of symptoms-adjusted | MDD PGS | 1.0497 | 0.0334 | 0.9831 – 1.1208 | 0.147 | 0.337 |
| | Number of episodes-adjusted | MDD PGS | 1.0486 | 0.0331 | 0.9827 – 1.1190 | 0.152 | 0.344 |
| | Kasper et al. definition of SAD | MDD PGS | 1.0819 | 0.0328 | 1.0144 – 1.1538 | **0.0165** | 0.103 |
| | Leave out calendar criterion of SAD | MDD PGS | 1.1232 | 0.0298 | 1.0595 – 1.1906 | **0.0000947** | **0.00161** |
| GSS | Longest episode-adjusted (NB) | MDD PGS | 1.0285 | 0.0064 | 1.0158 – 1.0415 | **0.0000105** | **0.0000444** |
| | Longest episode-adjusted (ZI) | MDD PGS | 1.0483 | 0.0398 | 0.9695 – 1.1334 | 0.237 | 1 |
| | Longest episode-adjusted (NB) | Longest episode | 1 | 0.0001 | 0.9999 – 1.0001 | 0.604 | 0.894 |
| | Longest episode-adjusted (ZI) | Longest episode | 1.0009 | 0.0002 | 1.0005 – 1.0012 | **0.000000851** | **0.00000965** |
| | Number of symptoms-adjusted (NB) | MDD PGS | 1.0204 | 0.0063 | 1.0079 – 1.0330 | **0.00128** | **0.00436** |
| | Number of symptoms-adjusted (ZI) | MDD PGS | 1.0645 | 0.0397 | 0.9849 – 1.1505 | 0.115 | 1 |
| | Number of symptoms-adjusted (NB) | Number of symptoms | 1.0979 | 0.0061 | 1.0848 – 1.1112 | **1.16e-52** | **3.94e-51** |
| | Number of symptoms-adjusted (ZI) | Number of symptoms | 0.9354 | 0.0363 | 0.8713 – 1.0043 | 0.0655 | 0.223 |
| | Number of episodes-adjusted (NB) | MDD PGS | 1.0249 | 0.0063 | 1.0124 – 1.0377 | **0.0000905** | **0.000385** |

| | | | | | | | |
|---|---|---|---|---|---|---|---|
| | Number of episodes-adjusted (ZI) | MDD PGS | 1.0357 | 0.0393 | 0.9589 – 1.1186 | 0.372 | 1 |
| | Number of episodes-adjusted (NB) | Number of episodes | 1.0121 | 0.0014 | 1.0093 – 1.0149 | **4.22e-17** | **7.18e-16** |
| | Number of episodes-adjusted (ZI) | Number of episodes | 1.0617 | 0.0093 | 1.0426 – 1.0812 | **0.000000000105** | **0.00000000179** |

*P*-adj = Multiple testing-adjusted P-value (Benjamini-Hochberg False Discovery Rate $P < 0.05$). SAD = seasonal affective disorder. GSS = general seasonality score. MDD = major depressive disorder. PGS = polygenic score. NB = count part of the zero-inflated negative binomial model. ZI = zero-inflated part of the negative binomial model. Bold indicates nominal significance ($P < 0.05$) or significance (FDR $P < 0.05$).
